# Association between genotypes of *ABCB1, ABCG2* and *CYP3A5* and the risk of atrial fibrillation

**DOI:** 10.1101/2024.07.09.24310172

**Authors:** Tzu-Yu Pan, Tzu-Yen Lin, Wei-Chung Tsai, Ming-Tsang Wu

**Affiliations:** Program in Environmental and Occupational Medicine, College of Medicine, Kaohsiung Medical University, Kaohsiung, Taiwan; Department of Public Health, College of Health Sciences, Kaohsiung Medical University, Kaohsiung, Taiwan; Division of Cardiology, Department of Internal Medicine, Kaohsiung Medical University Hospital, Kaohsiung, Taiwan; Faculty of Medicine, College of Medicine, Kaohsiung Medical University, Kaohsiung, Taiwan; Department of Family Medicine, Kaohsiung Medical University Hospital, Kaohsiung Medical University, Kaohsiung, Taiwan

**Keywords:** Single nucleotide polymorphisms, Atrial fibrillation, *ABCB1*, *ABCG2*, *CYP3A5*

## Abstract

**Background:** Atrial fibrillation (AF) is a prevalent clinical condition worldwide, with a high global incidence that significantly impacts disease burden and mortality rates. Single nucleotide polymorphisms in *ABCB1*, *ABCG2* and *CYP3A5* are common, but the clinical outcomes are poorly understood. This study examines the association between the genetic variations of *ABCB1, ABCG2* and *CYP3A5* and the risk of AF in a Taiwanese population.

**Methods:** This case-control study recruited 216 AF patients from two hospitals in Taiwan between 2021 and 2023. Control groups were matched by age (± one year), gender, and AF-related variables from the Taiwan Biobank. Logistic regression analyzed the association between three genetic variants and AF risk.

**Results:** A significant association was noted between *ABCG2 rs2231142* and AF risk. Those with *ABCG2 rs2231142 G/T* and *T/T* genotypes had a 1.91-fold (95% CI = 1.04-3.53) increased risk of AF compared to those with the G/G genotype. This association was particularly pronounced in males in those carrying *ABCG2 rs2231143 T/T* genotype having a 4.47-fold (95% CI = 1.02-19.67) increased risk after adjusting for covariates. There were no overall significant associations between AF risk and the polymorphisms of *ABCB1 rs4148738* and *rs1128503,* nor *CYP3A5 rs776746*.

**Conclusion:** A robust risk association between the *ABCG2 rs2231142 T allele* and AF in Asian populations, particularly in male adults, suggests that genetic testing for this polymorphism could be integrated into risk assessment models for AF.

## Introduction

Atrial fibrillation (AF) is recognized globally as a prevalent clinical condition that elevates the risk of systemic thromboembolism, heart failure, stroke, dementia and mortality, significantly impacting disease burden and mortality rates worldwide. ^1,2^ Current estimates indicate that approximately 33 million people are affected by AF globally, demonstrating its extensive reach and the urgency for comprehensive understanding and effective management strategies. ^3^ The prevalence of AF differs markedly between genders, with rates of 1.4% in males and 0.7% in females. Additionally, incidence rates are reported at 1.68 per 1000 males and 0.76 per 1000 females, highlighting a substantial gender disparity in AF occurrence. ^1,2^ Age also plays a crucial role, with the Global Burden of Disease Study 2017 (GBD 2017) revealing that AF prevalence increases with age—from 0.07% among those aged 30 to 34, to 8.18% for those aged 95 and above.^4^

Reports have also suggested potential disparities in the incidence and characteristics of AF across different racial and ethnic groups. ^5-7^ In Taiwan, heart disease, primarily driven by AF, ranks as the leading cause of death in individuals aged 65 and above and the second leading cause of death overall, following cancer, according to the 2021 national mortality statistics from the Ministry of Health and Welfare. ^2^ Extensive research has been directed towards unraveling the mechanisms underlying AF. Predominantly, AF is seen as a consequence of diseases such as hypertension, ischemic and structural heart disease, which contribute to the electrical and structural remodeling of the atria. ^8-10^ Despite the known risk factors such as advancing age, heart disease, elevated blood pressure and excessive alcohol consumption, a significant number of cases occur without identifiable risk factors, hinting at a potential genetic predisposition to AF. This gap in understanding limits the development of effective treatments and highlights the necessity for further investigation, particularly in the realm of precision medicine. ^2,11^

Over 60% of AF variability is attributed to genetic factors heightening AF susceptibility, influencing AF development and maintenance. ^12-14^ Genetic variations in the genes *ABCB1 (adenosine triphosphate (ATP) binding cassette subfamily B member 1), ABCG2 (ATP-binding cassette subfamily G member 2),* and *CYP3A5 (cytochrome P450 family 3 subfamily A member 5)*, responsible for encoding P-glycoprotein (P-gp), breast cancer resistance protein (BCRP) and cytochrome P450 proteins (CYP450) respectively, play a significant role in altering transporter function. These genetic variations impact crucial aspects of AF, including cardiac electrical conduction, ion channel function and structural remodeling. ^15,16^ Studies have found specific single nucleotide polymorphisms (SNPs) such as rs1128503 and rs4148738 in *ABCB1*, which modify P-gp expression; furthermore, *ABCG2 rs2231142 G>T* genotype has been linked to decreased BCRP protein expression, ^17,18^ while *CYP3A5* is crucial in metabolizing anticoagulants and stroke-prevention drugs, and this process significantly impacts the achievement of appropriate anticoagulation levels in patients with AF. ^19^

Understanding the relationship between these genetic variants and the risk of developing AF is essential as it can provide insight into underlying molecular mechanisms and potentially guide the development of targeted therapies in the era of precision medicine. ^20^ In recent years, precision medicine has garnered increasing attention, emphasizing the significant relationship between diseases and genes. ^21^ Each individual possesses a unique genome that can influence their disease susceptibility, treatment response and drug metabolism, ^22^ and through genetic testing and analysis, researchers can establish correlations between genetic variants and particular diseases. ^23^ This knowledge can predict an individual’s disease risk, facilitate accurate disease diagnosis, and aid in selecting the most suitable treatment options; ^24^ however, the current understanding of the relationship between these gene variants and the risk of AF remains unclear. In this study, we aimed to investigate the association between genetic variants of *ABCB1 (rs1128503, rs4148738), ABCG2 (rs2231142)* and *CYP3A5 (rs776746),* and the risk of AF.

## Materials and methods

### Study subjects

This is an ongoing case-control study. The case patients diagnosed as AF by board-certified cardiologists were recruited from Kaohsiung Medical University Hospital (KMUH) and Kaohsiung Municipal Ta-Tung Hospital (KMTTH) between January 2021 and January 2023. The study subjects included individuals aged 20-85 years and willing to cooperate with blood-draw procedures. Patients undergoing cancer treatment, those with impaired renal function (eGFR<15 ml/min/1.73 m^2^) and those who could not cooperate were excluded from the study. After the written informed consent was received, a designated researcher administered a standardized and structured questionnaire and reviewed the medical charts to collect age, gender, history of hypertension and diabetes and blood test data such as glycated hemoglobin (HbA1C), serum creatinine, triglyceride (TG) and low-density lipoprotein cholesterol (LDL)). A total of 216 AF patients were enrolled in the study period.

Potential healthy controls were selected from the Taiwan Biobank (TWB) cohort, a community-based cohort comprising healthy adults recruited from all over Taiwan. The TWB cohort, established in 2005, aimed to collect genetic and medical information from 200,000 ethnic Taiwanese individuals aged 20 years or above without a history of cancer, where this cohort underwent comprehensive data collection including questionnaires, physical examinations and biochemical tests on blood and urine samples, and has continued to collect biological samples to-date, including DNA, blood plasma and urine from study subjects, which can be provided to researchers in Taiwan for study purposes at a minimal cost. As of 2020, the TWB cohort had enrolled 122,068 potential study adults aged 30-75 years with baseline data. The Institutional Review Board (IRB) of KMUH approved this study.

Among the 216 AF patients, 185 subjects have provided blood samples for genotyping results. In addition, in order to match the AF patients of similar age with healthy controls from TWB cohort, we further excluded 44 patients aged over 76 years and left 141 eligible cases in the study (**Figure 1**). Then, the case patients were matched with the controls (1: 1 ratio) based on age within a range of ±1 year and the same gender from TWB cohort with specific genotypes of *ABCB1 (rs1128503, rs4148738), ABCG2 (rs2231142),* and *CYP3A5 (rs776746)* (n = 108,829). The final study subjects were 141 AF cases and 141 healthy controls.

**Figure 1.**
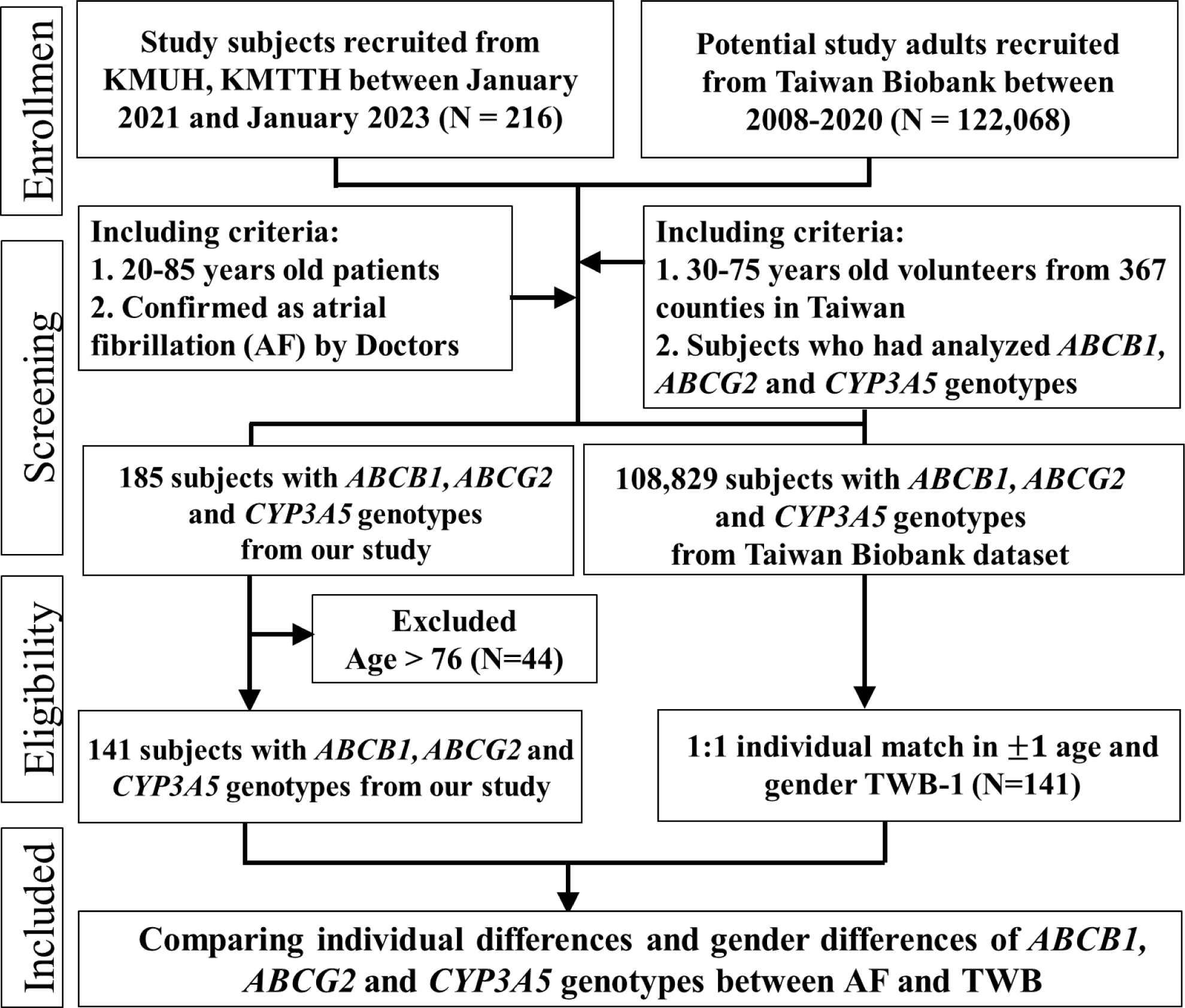
Flow chart comparing the genetic risk of AF in our study patients and Taiwan biobank’s healthy subjects.

### Genotyping of ABCB1 (rs1128503, rs4148738), ABCG2 (rs2231142) and CYP3A5 (rs776746) **polymorphisms from the case study group**

The QIAamp DNA Mini Kit (Qiagen, Germany) was used to extract genomic DNA from peripheral blood samples collected from patients in the case study group, following the manufacturer’s instructions, while TaqMan probes designed explicitly for standardized SNPs assays (Thermo Fisher Scientific, Inc) were employed to assess single nucleotide polymorphic variations in the *ABCB1* (rs1128503, rs4148738), *ABCG2* (rs2231142) and *CYP3A5* (rs776746) genes. Probe IDs and SNPs regions are listed in **Table S1**.

For SNPs genotyping, a reaction mixture containing 10 μL was prepared, containing 10 ng of genomic DNA, 2X TaqMan® Master Mix, 20X Assay Working Stock probe and nuclease-free water. A negative control (no template) was included in the experiment. The cycling conditions adhered to the manufacturer’s standard protocol as recommended where real-time PCR was conducted on all samples using the QuantStudio 3 instrument (Applied Biosystems) and QuantStudio™ Design & Analysis Software was employed to analyze the data and identify homozygous and heterozygous alleles. To ensure the reliability of the genotyping assays, approximately 9.4% of patients (n = 15/141) were randomly selected for the genotyping of each polymorphism; this additional step being performed to validate the consistency and accuracy of the genotyping results. The reaction of PCR for direct sequencing is shown in **Supplemental Material and Table S2** that was also performed to confirm the genotyping results (**Figure S1**). For TWB cohorts, we extracted the genotype information of *ABCB1* (rs1128503, rs4148738), *ABCG2* (rs2231142), and *CYP3A5* (rs776746) genes from Genome-Wide Genotyping: Affymetrix Axiom TWB and TWB 2.0 database, in which genotype analysis was either from the Axiom Genome-Wide TWB array or Axiom Genome-Wide TWB 2.0 array. ^25^

### Statistical analysis

The frequencies of *ABCB1 (rs1128503), ABCG2 (rs2231142)* and *CYP3A5 (rs776746)* genotypes were assessed for Hardy-Weinberg disequilibrium in the TWB-1 control group (n = 141). A comparison of demographic characteristics and genotypes between the case group (n = 141) and the TWB-1 control group (n = 141) was performed. Logistic regression models were employed to investigate the impact of *ABCB1 (rs1128503, rs4148738), ABCG2 (rs2231142),* and *CYP3A5 (rs776746)* genotypes on AF risk, both before and after adjusting for covariates, including age (continuous variables), gender, diabetes, hypertension, HbA1C, serum creatinine, TG and LDL. Odds ratios (OR), confidence intervals (CI) and p-values were calculated and presented. If both *ABCB1 (rs1128503, rs4148738), ABCG2 (rs2231142)* and *CYP3A5 (rs776746)* genotypes showed significant differences between case patients and controls, the combined effect of both genes was further examined. These analyses were also stratified by gender. All statistical analyses were conducted using IBM SPSS Statistics 25 software.

## Results

### Characteristics of the Study Population

The demographic characteristics of 141 AF patients and the 141 TWB-1 controls are shown in **Table 1**. Among these 141 AF patients, 48 (34.0%) were female and 93 (66.0%) were male, while the percentages of gender, age group (40-60 and > 60 years) and educational levels were identical in AF patients and TWB-1 controls. Additionally, between the two groups of AF patients and TWB-1 controls, the aforementioned variables (gender, age groups and other variables including diabetes, hypertension, HbA1c, serum creatinine, TG and LDL) were not significantly different (all p > 0.05). Of the 141 AF patients, 24.8% had diabetes and 35.5% had hypertension; additionally, the HbA1c level (6.2%) was higher than the normal range of 4.0-5.6%.

**Table 1.**
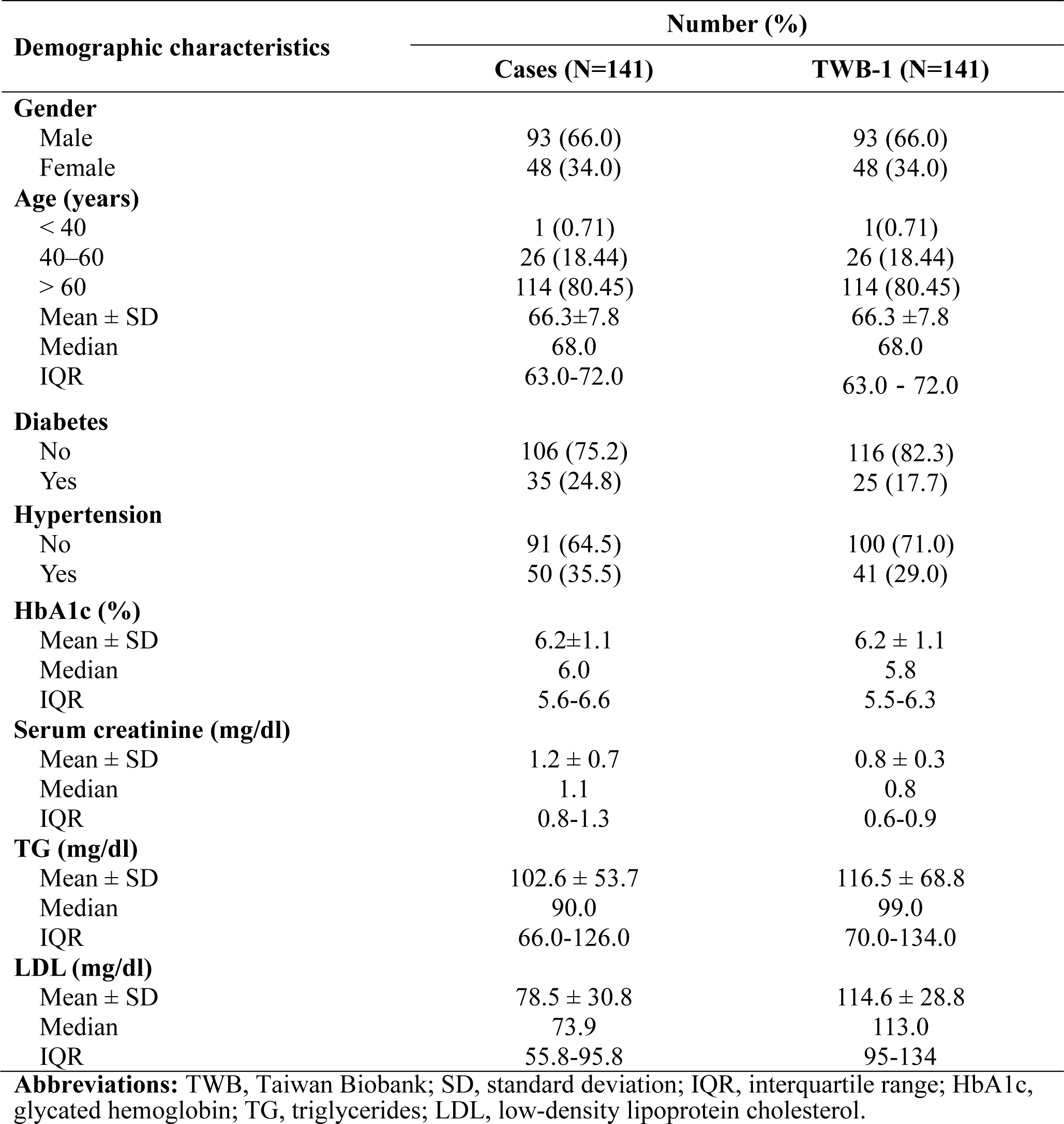
Demographic characteristics of atrial fibrillation and their comparison groups.

| Demographic characteristics | Number (%) |  |
| --- | --- | --- |
|  | Cases (N=141) | TWB-1 (N=141) |
| **Gender** |  |  |
| Male | 93 (66.0) | 93 (66.0) |
| Female | 48 (34.0) | 48 (34.0) |
| **Age (years)** |  |  |
| < 40 | 1 (0.71) | 1(0.71) |
| 40–60 | 26 (18.44) | 26 (18.44) |
| > 60 | 114 (80.45) | 114 (80.45) |
| Mean ± SD | 66.3±7.8 | 66.3 ±7.8 |
| Median | 68.0 | 68.0 |
| IQR | 63.0-72.0 | 63.0 - 72.0 |
| **Diabetes** |  |  |
| No | 106 (75.2) | 116 (82.3) |
| Yes | 35 (24.8) | 25 (17.7) |
| **Hypertension** |  |  |
| No | 91 (64.5) | 100 (71.0) |
| Yes | 50 (35.5) | 41 (29.0) |
| **HbA1c (%)** |  |  |
| Mean ± SD | 6.2±1.1 | 6.2 ± 1.1 |
| Median | 6.0 | 5.8 |
| IQR | 5.6-6.6 | 5.5-6.3 |
| **Serum creatinine (mg/dl)** |  |  |
| Mean ± SD | 1.2 ± 0.7 | 0.8 ± 0.3 |
| Median | 1.1 | 0.8 |
| IQR | 0.8-1.3 | 0.6-0.9 |
| **TG (mg/dl)** |  |  |
| Mean ± SD | 102.6 ± 53.7 | 116.5 ± 68.8 |
| Median | 90.0 | 99.0 |
| IQR | 66.0-126.0 | 70.0-134.0 |
| **LDL (mg/dl)** |  |  |
| Mean ± SD | 78.5 ± 30.8 | 114.6 ± 28.8 |
| Median | 73.9 | 113.0 |
| IQR | 55.8-95.8 | 95-134 |

**Abbreviations:** TWB, Taiwan Biobank; SD, standard deviation; IQR, interquartile range; HbA1c, glycated hemoglobin; TG, triglycerides; LDL, low-density lipoprotein cholesterol.

### Allele frequencies in TWB cohort

The prevalence of *ABCB1 rs1128503 A/A, A/G,* and *G/G* was 41.8%, 46.8% and 11.3% respectively, for TWB-1 (n = 141). For *ABCB1 rs4148738 T/T, C/T,* and *C/C*, the prevalence was 34.0%, 41.8% and 24.1% respectively, for TWB-1. *ABCG2 rs2231142 G/G, G/T,* and *T/T* were 59.6%, 33.3% and 7.1% respectively, in TWB-1. *CYP3A5 rs776746 C/C, T/C,* and *T/T* were 46.1%, 44.0% and 9.9% respectively, in TWB-1 (**Table S3**). All frequencies of *ABCB1 (rs1128503, rs4148738), ABCG2 (rs2231142),* and *CYP3A5 (rs776746)* genotypes in TWB-1 were consistent with the statistics of the Hardy-Weinberg equilibrium, which were insignificant (p > 0.05) (**Table S3**).

### Association of ABCB1, ABCG2 and CYP3A5 polymorphisms

**Table 2** shows the association of *ABCB1 (rs1128503, rs4148738), ABCG2 (rs2231142),* and *CYP3A5 (rs776746)* genotypes with AF risk before and after adjusting for other covariates (including diabetes, hypertension, HbA1c, serum creatinine, TG and LDL). Genotypes of *ABCG2 rs2231142 G/T* and *T/T* exhibited a significant risk of developing AF compared to *G/G* genotypes after adjusting for other covariates (AOR = 1.912, 95% CI = 1.035-3.53), although no significant difference was observed in the other three SNPs after adjustment.

**Table 2.**
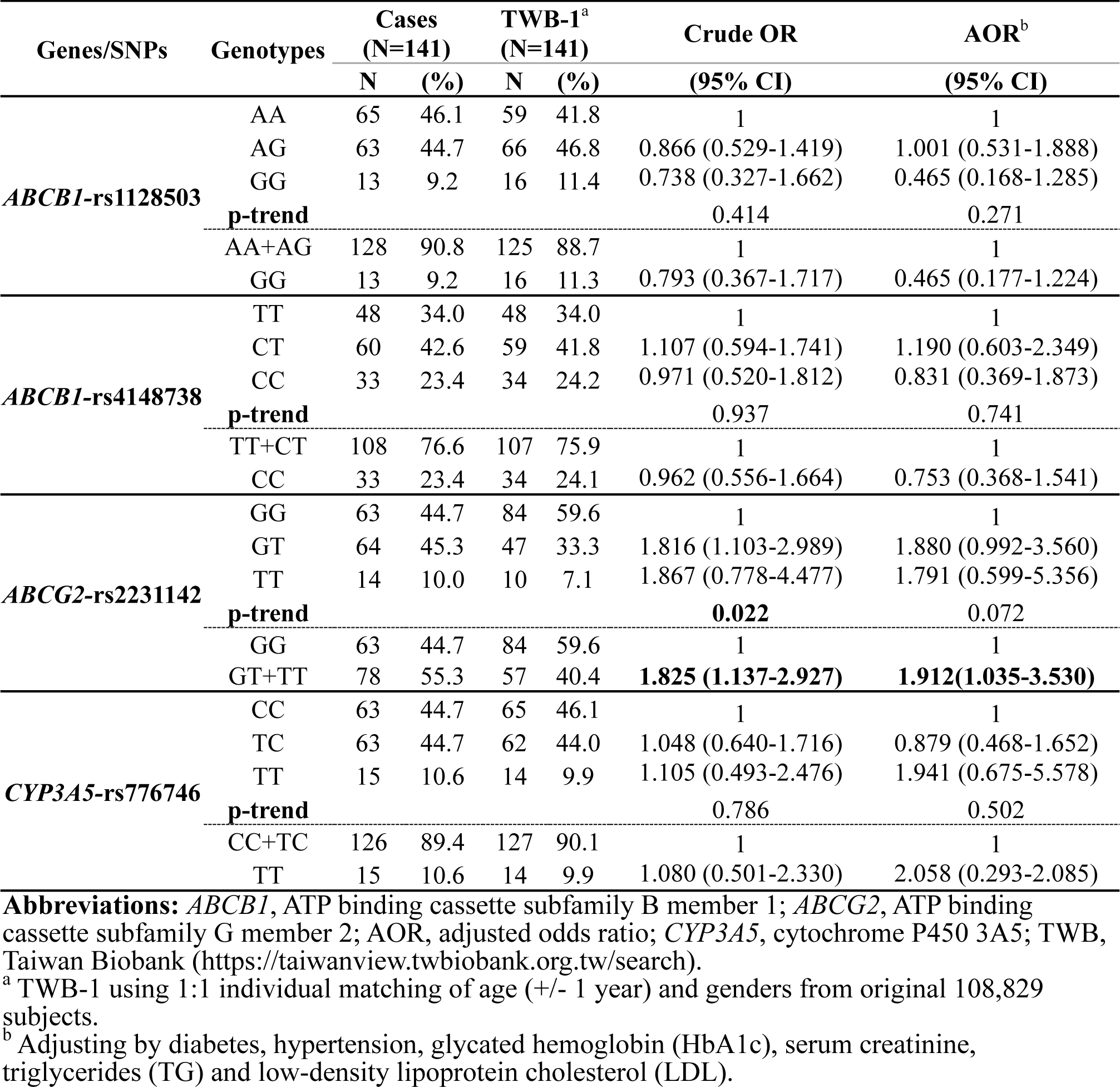
Distribution of *ABCB1*, *ABCG2*, and *CYP3A5* genotypes and odds ratios among atrial fibrillation cases and Taiwan biobank controls.

Categorized by sex, the significant effect of *ABCG2 rs2231143* remained present in males but not females, likely due to the small sample size of female AF patients (n = 48) (**Table 3**). Males with the combined genotypes of *ABCG2 rs2231143 G/T* and *T/T* had a significant risk of AF when compared with males carrying *ABCG2 rs2231143 G/G* genotype (crude OR=2.005, 95% CI=1.118-3.596; AOR=2.179, 95% CI=0.894-5.307). There was also a trend toward statistical significance for the association between male subjects and AF (p = 0.021 and 0.041 before and after adjusting for other covariates), suggesting a potential risk factor associated with this haplotype in males.

**Table 3.**
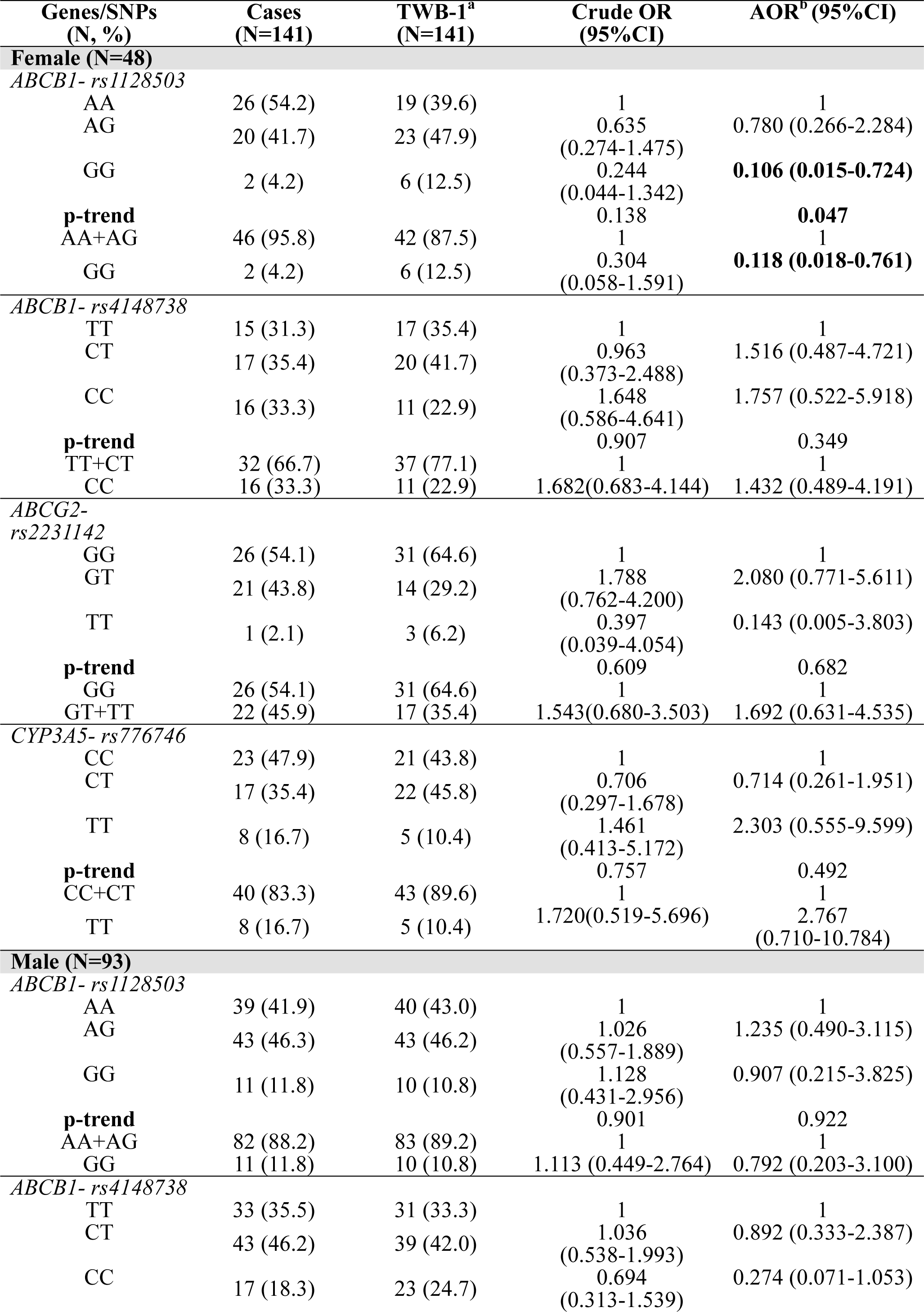
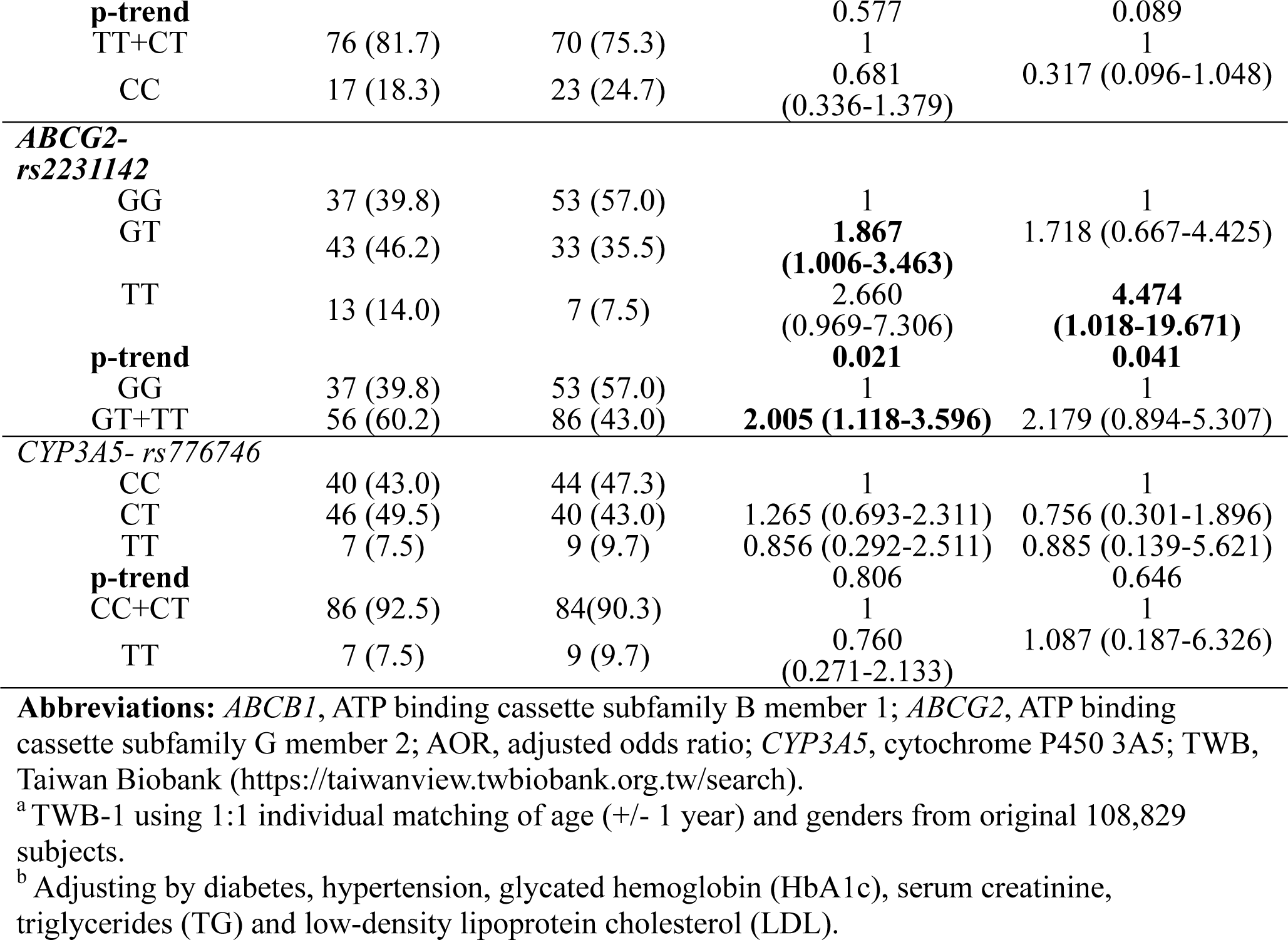
Logistic regression analysis with risk estimates of *ABCB1, ABCG2,* and *CYP3A5* genotypes in Taiwanese female and male atrial fibrillation patients.

| Genes/SNPs<br>(N, %) | Cases<br>(N=141) | TWB-1<sup>a</sup><br>(N=141) | Crude OR<br>(95%CI) | AOR<sup>b</sup> (95%CI) |
| --- | --- | --- | --- | --- |
| **Female (N=48)** |  |  |  |  |
| *ABCB1*- *rs1128503* |  |  |  |  |
| AA | 26 (54.2) | 19 (39.6) | 1 | 1 |
| AG | 20 (41.7) | 23 (47.9) | 0.635<br>(0.274-1.475) | 0.780 (0.266-2.284) |
| GG | 2 (4.2) | 6 (12.5) | 0.244<br>(0.044-1.342) | **0.106 (0.015-0.724)** |
| **p-trend** |  |  | 0.138 | **0.047** |
| AA+AG | 46 (95.8) | 42 (87.5) | 1 | 1 |
| GG | 2 (4.2) | 6 (12.5) | 0.304<br>(0.058-1.591) | **0.118 (0.018-0.761)** |
| *ABCB1*- *rs4148738* |  |  |  |  |
| TT | 15 (31.3) | 17 (35.4) | 1 | 1 |
| CT | 17 (35.4) | 20 (41.7) | 0.963<br>(0.373-2.488) | 1.516 (0.487-4.721) |
| CC | 16 (33.3) | 11 (22.9) | 1.648<br>(0.586-4.641) | 1.757 (0.522-5.918) |
| **p-trend** |  |  | 0.907 | 0.349 |
| TT+CT | 32 (66.7) | 37 (77.1) | 1 | 1 |
| CC | 16 (33.3) | 11 (22.9) | 1.682(0.683-4.144) | 1.432 (0.489-4.191) |
| *ABCG2*-<br>*rs2231142* |  |  |  |  |
| GG | 26 (54.1) | 31 (64.6) | 1 | 1 |
| GT | 21 (43.8) | 14 (29.2) | 1.788<br>(0.762-4.200) | 2.080 (0.771-5.611) |
| TT | 1 (2.1) | 3 (6.2) | 0.397<br>(0.039-4.054) | 0.143 (0.005-3.803) |
| **p-trend** |  |  | 0.609 | 0.682 |
| GG | 26 (54.1) | 31 (64.6) | 1 | 1 |
| GT+TT | 22 (45.9) | 17 (35.4) | 1.543(0.680-3.503) | 1.692 (0.631-4.535) |
| *CYP3A5*- *rs776746* |  |  |  |  |
| CC | 23 (47.9) | 21 (43.8) | 1 | 1 |
| CT | 17 (35.4) | 22 (45.8) | 0.706<br>(0.297-1.678) | 0.714 (0.261-1.951) |
| TT | 8 (16.7) | 5 (10.4) | 1.461<br>(0.413-5.172) | 2.303 (0.555-9.599) |
| **p-trend** |  |  | 0.757 | 0.492 |
| CC+CT | 40 (83.3) | 43 (89.6) | 1 | 1 |
| TT | 8 (16.7) | 5 (10.4) | 1.720(0.519-5.696) | 2.767<br>(0.710-10.784) |
| **Male (N=93)** |  |  |  |  |
| *ABCB1*- *rs1128503* |  |  |  |  |
| AA | 39 (41.9) | 40 (43.0) | 1 | 1 |
| AG | 43 (46.3) | 43 (46.2) | 1.026<br>(0.557-1.889) | 1.235 (0.490-3.115) |
| GG | 11 (11.8) | 10 (10.8) | 1.128<br>(0.431-2.956) | 0.907 (0.215-3.825) |
| **p-trend** |  |  | 0.901 | 0.922 |
| AA+AG | 82 (88.2) | 83 (89.2) | 1 | 1 |
| GG | 11 (11.8) | 10 (10.8) | 1.113 (0.449-2.764) | 0.792 (0.203-3.100) |
| *ABCB1*- *rs4148738* |  |  |  |  |
| TT | 33 (35.5) | 31 (33.3) | 1 | 1 |
| CT | 43 (46.2) | 39 (42.0) | 1.036<br>(0.538-1.993) | 0.892 (0.333-2.387) |
| CC | 17 (18.3) | 23 (24.7) | 0.694<br>(0.313-1.539) | 0.274 (0.071-1.053) |
| **p-trend** |  |  | 0.577 | 0.089 |
| TT+CT | 76 (81.7) | 70 (75.3) | 1 | 1 |
| CC | 17 (18.3) | 23 (24.7) | 0.681<br>(0.336-1.379) | 0.317 (0.096-1.048) |
| **ABCG2-**<br>**rs2231142** |  |  |  |  |
| GG | 37 (39.8) | 53 (57.0) | 1 | 1 |
| GT | 43 (46.2) | 33 (35.5) | **1.867**<br>**(1.006-3.463)** | 1.718 (0.667-4.425) |
| TT | 13 (14.0) | 7 (7.5) | 2.660<br>(0.969-7.306) | **4.474**<br>**(1.018-19.671)** |
| **p-trend** |  |  | **0.021** | **0.041** |
| GG | 37 (39.8) | 53 (57.0) | 1 | 1 |
| GT+TT | 56 (60.2) | 86 (43.0) | **2.005 (1.118-3.596)** | 2.179 (0.894-5.307) |
| **CYP3A5- rs776746** |  |  |  |  |
| CC | 40 (43.0) | 44 (47.3) | 1 | 1 |
| CT | 46 (49.5) | 40 (43.0) | 1.265 (0.693-2.311) | 0.756 (0.301-1.896) |
| TT | 7 (7.5) | 9 (9.7) | 0.856 (0.292-2.511) | 0.885 (0.139-5.621) |
| **p-trend** |  |  | 0.806 | 0.646 |
| CC+CT | 86 (92.5) | 84(90.3) | 1 | 1 |
| TT | 7 (7.5) | 9 (9.7) | 0.760<br>(0.271-2.133) | 1.087 (0.187-6.326) |

**Abbreviations:** *ABCB1*, ATP binding cassette subfamily B member 1; *ABCG2*, ATP binding cassette subfamily G member 2; AOR, adjusted odds ratio; *CYP3A5*, cytochrome P450 3A5; TWB, Taiwan Biobank (https://taiwanview.twbiobank.org.tw/search).
<sup>a</sup> TWB-1 using 1:1 individual matching of age (+/- 1 year) and genders from original 108,829 subjects.
<sup>b</sup> Adjusting by diabetes, hypertension, glycated hemoglobin (HbA1c), serum creatinine, triglycerides (TG) and low-density lipoprotein cholesterol (LDL).

## Discussion

This hospital-based case-control study underscores the importance of the *ABCG2 rs2231142 G/T* and *T/T* genotypes, which exhibit a higher risk of developing AF than those with the G/G genotype, particularly among male patients. However, no associations were identified between the polymorphisms of *ABCB1 rs4148738* and *rs1128503,* and *CYP3A5 rs776746* genotypes and AF risk.

*ABCG2* belongs to a superfamily of 48 human ATP-binding cassette transporters, which typically facilitate the transport of various substrates and rely on ATP-binding for activation. The *ABCG2 rs2231142* variant results in a substitution of glutamine with lysine (Q141K) in exon 5. Previous studies have reported that individuals carrying the *ABCG2 rs2231142* variant (*G/T* and *T/T*) exhibit significantly lower levels of ABCG2 protein within red blood cells as well as reduced ABCG2 adenosine triphosphatase (ATPase) activity compared to those with the *G/G* genotype.

The presence of the *ABCG2 rs2231142 T* allele leads to an approximately 50% reduction in ABCG2 protein expression. ^26^ Zhang et al. found that variations in the *ABCG2 rs2231142* variant could impact the serum levels of pro-inflammatory and pro-angiogenic markers in chronic inflammatory arterial conditions, hinting at a correlation between inflammation and the *ABCG2 rs2231142* variant. ^27^ These findings suggest that *ABCG2 rs2231142* could play a crucial physiological role as an excreter of environmental and endogenous toxins, and it might influence the multidrug transporter function in human erythrocytes, corresponding to pharmacologically relevant genetic variations. ^28^ Furthermore, recent studies have emphasized that the pharmacogenetic implications of these findings could connote the efficacy of direct oral anticoagulants (DOACs) being enhanced, as genetic variants influence drug metabolism and response. ^29,30^

In recent years, DOACs such as apixaban, dabigatran, edoxaban and rivaroxaban have replaced vitamin K antagonists for treating AF due to their ease of use and better effectiveness. In a rat model of pharmacokinetic interaction, almonertinib significantly increased systemic exposure to apixaban and rivaroxaban by inhibiting *ABCB1* and *ABCG2.* ^31^ Previous studies have shown variable results regarding the relationship between genotypes and DOACs in AF patients. Gulilat et al. reported that the *ABCG2 rs2231142 T* allele predicted higher concentrations of apixaban in Caucasian AF patients, ^32^ but was associated with a lower concentration and lower oral clearance of apixaban in Japanese AF patients. ^33,34^ Additionally, no correlations were observed between the *ABCG2 rs2231142* and rivaroxaban concentrations in Japanese AF patients. ^35^ Nevertheless, we observed a significant increase in the risk of AF associated with the *ABCG2 rs2231142 T* allele in patients treated with rivaroxaban (**Table S4**), indicating that this SNPs might affect the drug transporters and metabolism of DOACs, making additional studies necessary to clarify the role of this SNPs in DOACs pharmacokinetic interactions.

Previous studies have shown that the loss-of-function *ABCG2 rs2231142 T* allele is associated with hyperuricemia, which might contribute to vascular damage and cardiovascular diseases such as atrial fibrillation, coronary artery disease, heart failure and stroke. ^36-38^ Similar observations were noted in an earlier prospective cohort study conducted in Taiwan, where hyperuricemia emerged as an independent risk factor for mortality from all causes, total cardiovascular disease and ischemic stroke. ^39^ In other studies, it is also suggested that the effect size of the *ABCG2 rs2231142* is more significant in males. ^36,40,41^ Our current data showed a similar interaction, as we found a significant association between the *rs2231142 T* allele and AF risk in males, which was consistent with these Asian population studies, ^36-41^ and although previous studies have demonstrated that estrogen levels could affect *ABCG2* expression, ^42,43^ this might explain why we observed the association with atrial fibrillation risk predominantly in males rather than females.

Currently, no studies have examined the AF risk linked to the *ABCG2 rs2231142* genotype, but research on other SNPs regarding AF risk has been conducted. Genetic factors contributing to AF are gaining more attention, with estimates of heritability reaching 22% in the UK Biobank and 62% in a twin study conducted in Denmark. ^14,44^ Daniel et al. were the first to report the Genome-wide association studies (GWAS) for AF, where the upstream region of the transcription factor gene *PITX2* on 4q25 was significantly associated with AF. ^45^ Several other genetic loci related to AF have been identified, including variants in or near *ZFHX3 (Zinc Finger Homeobox 3), PITX2 (paired like homeodomain 2)*, and *MYOZ1 (myozenin 1)*, which show the most associations. ^46^

Our research further highlights the significant relationship between the *ABCG2 rs2231142 T* allele and increased AF risk, particularly in males. This suggests that integrating genetic testing for this polymorphism into risk assessment models for AF could enhance precision in AF management strategies, especially in populations with a high prevalence of this risk allele. Past studies have primarily focused on pharmacogenomics or the risk of cardiovascular and hyperuricemia. ^36-41^ The association between polymorphisms of *ABG2 rs2231142* genotypes and AF subjects might result from the influence of hyperuricemia, obesity, hypertension and dyslipidemia, as well as sex differences caused by estrogen levels, or both. ^47^ In the study conducted by Doshi et al., ^48^ *ABCG2* expression was examined in male obesity mice. The findings indicated elevated levels of *ABCG2* protein, suggesting a potential association between increased *ABCG2* expression and obesity-related AF, although the mechanism of the influence of the *ABCG2 rs2231142* locus on the risk effect of developing AF still needs to be investigated.

**Table S5** shows the similar variant allele frequencies of *ABCB1, ABCG2* and *CYP3A5* genotypes in East Asians from different databases and from the datasets of TWB, suggesting the robustness of target genotype frequencies in the public-domain databases. The 1000 Genomes Project (1000Genomes) and the Genome Aggregation Database (gnomAD) are public websites that respectively encompass data from more than 2,500 individuals representing global populations and aggregate over 800,000 exome and whole-genome sequences from seven ethnic groups, sharing summary data with the scientific community (https://www.internationalgenome.org/ and https://reurl.cc/RWnZzg, accessed on 2024/4/18). The TWB is an ongoing prospective study that involves over 200,000 individuals aged 20–70 in Taiwan.

The distributions of genotype and allelic frequencies in Taiwan and other populations such as American, European, African, South Asian, Central Asian and East Asian are summarized in **Table S6**. The distributions of allelic frequencies in other populations were taken from the database of NCBI (https://www.ncbi.nlm.nih.gov/, assessed on 2024/4/18). The *ABCB1 rs1128503 G* allele and *rs4148738 T* allele have a higher prevalence among Africans, with frequencies of 80% and 77% respectively, while the *ABCG2 T* allele is more common in American, East Asian and Taiwanese populations consistent with the findings reported by Alrajeh et al. ^49^ Their reports noted that the frequency of the allele associated with reduced *ABCG2* function is exceptionally high among Asians while the frequency of the *CYP3A5 C* allele is lower in African populations, with a frequency of 31%; interestingly, the prevalence of the *ABCG2 T* allele is higher in East Asian, Taiwanese and American populations (29.8%, 31.5% and 22.5%, respectively) compared to European, South Asian and African populations (9.8%, 9% and 2.6%, respectively). These differences may be attributed to factors such as database sources, study designs and ethnicities.

## Conclusion

In conclusion, the findings suggest a strong association between the *ABCG2 rs2231142 T* allele and a higher risk of AF among Taiwanese male subjects, with a 4.5-fold higher risk compared to controls, and a 1.9-fold risk when combining *ABCG2* genotypes in the overall population. However, no significant associations were identified between the polymorphisms of *ABCB1 rs4148738* and *rs1128503*, as well as *CYP3A5 rs776746* genotypes and AF risk. Further studies with larger sample sizes are necessary to investigate the interaction of the *ABCG2 rs2231142* genotype with AF risk.

## Data Availability

The data are not publicly available due to the nature of information that could compromise the privacy of research participants.

## Nonstandard Abbreviations and Acronyms

1000Genomes: 1000 Genomes Project
AF: Atrial fibrillation
ABCB1: Adenosine triphosphate binding cassette subfamily B member 1
ABCG2: ATP-binding cassette subfamily G member 2
ATPase: Adenosine triphosphatase
BCRP: Breast cancer resistance protein
CYP3A5: Cytochrome P450 family 3 subfamily A member 5
CYP450: Cytochrome P450 proteins
DOACs: Direct oral anticoagulants
GBD 2017: Global Burden of Disease Study 2017
gnomAD: Genome Aggregation Database
GWAS: Genome-Wide Association Studies
HbA1C: Glycated hemoglobin
IRB: Institutional Review Board
KMUH: Kaohsiung Medical University Hospital
KMTTH: Kaohsiung Municipal Ta-Tung Hospital
LDL: Low-density lipoprotein cholesterol
MYOZ1: Myozenin 1
P-gp: P-glycoprotein
PITX2: Paired like homeodomain 2
SNPs: Single Nucleotide Polymorphisms
TG: Triglyceride
TWB: Taiwan Biobank
ZFHX3: Zinc finger homeobox 3

## Acknowledgment

Not applicable.

## Authors’ contributions

**Tzu-Yen Lin** was responsible for Methodology, Validation, Investigation, Data curation; **Tzu-Yu Pan** was responsible for Methodology, Validation, Formal analysis, Resources, Data curation, Writing – original draft and Writing-Reviewing and Editing; **Wei-Chung Tsai** were responsible for Data Curation and Resources, Writing-Review and editing; **Ming-Tsang Wu** was responsible for Conceptualization, Writing-Review and editing, Supervision, Project administration, Funding acquisition.

## Ethics approval and consent to participate

The study design was approved by the institutional review board of Kaohsiung Medical University Hospital (KMUHIRB-G(I)-20200040).

## Supplemental Material

Supplemental Method

Tables S1–S6

Figure S1

References ^50^

## References

1. Chien K-L, Su T-C, Hsu H-C, Chang W-T, Chen P-C, Chen M-F, Lee Y-T. Atrial fibrillation prevalence, incidence and risk of stroke and all-cause death among Chinese. International journal of cardiology. 2010;139:173–180. doi: 10.1016/j.ijcard.2008.10.045

2. Andersen JH, Andreasen L, Olesen MS. Atrial fibrillation—a complex polygenetic disease. European Journal of Human Genetics. 2021;29:1051–1060. doi: 10.1038/s41431-020-00784-8

3. Schnabel RB, Yin X, Gona P, Larson MG, Beiser AS, McManus DD, Newton-Cheh C, Lubitz SA, Magnani JW, Ellinor PT. 50 year trends in atrial fibrillation prevalence, incidence, risk factors, and mortality in the Framingham Heart Study: a cohort study. The Lancet. 2015;386:154–162. doi: 10.1016/S0140-6736(14)61774-8

4. Melki S, Salem B, Soulimane A, Serhier Z, Dahdi S. Profile and evolution of the Global Burden of Morbidity in the Maghreb (Tunisia, Morocco, Algeria). The Triple burden of morbidity. La Tunisie Medicale. 2018;96:760–773.

5. Zhang J, Johnsen SP, Guo Y, Lip GY. Epidemiology of atrial fibrillation: geographic/ecological risk factors, age, sex, genetics. Cardiac electrophysiology clinics. 2021;13:1–23. doi: 10.1016/j.ccep.2020.10.010

6. Nanda A, Kabra R. Racial differences in atrial fibrillation epidemiology, management, and outcomes. Current Treatment Options in Cardiovascular Medicine. 2019;21:1–11. doi: 10.1007/s11936-019-0793-5

7. Tamirisa KP, Al-Khatib SM, Mohanty S, Han JK, Natale A, Gupta D, Russo AM, Al-Ahmad A, Gillis AM, Thomas KL. Racial and ethnic differences in the management of atrial fibrillation. CJC open. 2021;3:S137–S148. doi: 10.1016/j.cjco.2021.09.004

8. Čihák R, Haman L, Táborský M. 2016 ESC Guidelines for the management of atrial fibrillation developed in collaboration with EACTS. Cor et vasa. 2016;6:e636–e683. doi: 10.1093/eurheartj/ehw210

9. Gudbjartsson DF, Holm H, Sulem P, Masson G, Oddsson A, Magnusson OT, Saemundsdottir J, Helgadottir HT, Helgason H, Johannsdottir H. A frameshift deletion in the sarcomere gene MYL4 causes early-onset familial atrial fibrillation. European heart journal. 2016:ehw379. doi: 10.1093/eurheartj/ehw379

10. Nattel S. Molecular and cellular mechanisms of atrial fibrosis in atrial fibrillation. JACC: Clinical Electrophysiology. 2017;3:425–435. doi: 10.1016/j.jacep.2017.03.002

11. Baturina O, Chashkina M, Andreev D, Mirzaev K, Bykova A, Suvorov A, Yeryshova D, Suchkova S, Sychev D, Syrkin A. Pharmacokinetic and Pharmacogenetic Predictors of Major Bleeding Events in Patients with an Acute Coronary Syndrome and Atrial Fibrillation Receiving Combined Antithrombotic Therapy. Journal of Personalized Medicine. 2023;13:1371. doi: 10.3390/jpm13091371

12. Chugh SS, Havmoeller R, Narayanan K, Singh D, Rienstra M, Benjamin EJ, Gillum RF, Kim Y-H, McAnulty Jr JH, Zheng Z-J. Worldwide epidemiology of atrial fibrillation: a Global Burden of Disease 2010 Study. Circulation. 2014;129:837–847. doi: 10.1161/CIRCULATIONAHA.113.005119

13. Ishikawa K, Hajjar RJ. Current methods in cardiac gene therapy: overview. Cardiac Gene Therapy: Methods and Protocols. 2017:3–14. doi: 10.1007/978-1-4939-6588-5_1

14. Christophersen IE, Ravn LS, Budtz-Joergensen E, Skytthe A, Haunsoe S, Svendsen JH, Christensen K. Familial aggregation of atrial fibrillation: a study in Danish twins. Circulation: Arrhythmia and Electrophysiology. 2009;2:378–383. doi: 10.1161/CIRCEP.108.786665

15. Lubitz SA, Lunetta KL, Lin H, Arking DE, Trompet S, Li G, Krijthe BP, Chasman DI, Barnard J, Kleber ME. Novel genetic markers associate with atrial fibrillation risk in Europeans and Japanese. Journal of the American College of Cardiology. 2014;63:1200–1210. doi: 10.1016/j.jacc.2013.12.015

16. Christophersen IE, Ellinor PT. Genetics of atrial fibrillation: from families to genomes. Journal of human genetics. 2016;61:61–70. doi: 10.1038/jhg.2015.44

17. Gong IY, Mansell SE, Kim RB. Absence of both MDR 1 (ABCB 1) and Breast Cancer Resistance Protein (ABCG 2) Transporters Significantly Alters Rivaroxaban Disposition and Central Nervous System Entry. Basic & clinical pharmacology & toxicology. 2013;112:164–170. doi: 10.1111/bcpt.12005

18. Mizuno T, Fukudo M, Fukuda T, Terada T, Dong M, Kamba T, Yamasaki T, Ogawa O, Katsura T, Inui K-i. The effect of ABCG2 genotype on the population pharmacokinetics of sunitinib in patients with renal cell carcinoma. Therapeutic drug monitoring. 2014;36:310–316. doi: 10.1097/FTD.0000000000000025

19. Sychev DA, Sokolov AV, Reshetko OV, Fisenko VP, Sychev IN, Grishina EA, Bochkov PO, Shevchenko RV, Abdullaev SP, Denisenko NP. Influence of ABCB1, CYP3A5 and CYP3A4 gene polymorphisms on prothrombin time and the residual equilibrium concentration of rivaroxaban in patients with non-valvular atrial fibrillation in real clinical practice. Pharmacogenetics and Genomics. 2022;32:301-307. doi: 10.1097/FPC.0000000000000483

20. Sinner MF, Ellinor PT, Meitinger T, Benjamin EJ, Kääb S. Genome-wide association studies of atrial fibrillation: past, present, and future. Cardiovascular research. 2011;89:701–709. doi: 10.1093/cvr/cvr001

21. König IR, Fuchs O, Hansen G, von Mutius E, Kopp MV. What is precision medicine? European respiratory journal. 2017;50. doi: 10.1183/13993003.00391-2017

22. Ramaswami R, Bayer R, Galea S. Precision medicine from a public health perspective. Annual Review of Public Health. 2018;39:153–168. doi: 10.1146/annurev-publhealth-040617-014158

23. Robinson PN. Deep phenotyping for precision medicine. Human mutation. 2012;33:777–780. doi: 10.1002/humu.22080

24. McGrath S, Ghersi D. Building towards precision medicine: empowering medical professionals for the next revolution. BMC medical genomics. 2016;9:1–6. doi: 10.1186/s12920-016-0183-8

25. Feng Y-CA, Chen C-Y, Chen T-T, Kuo P-H, Hsu Y-H, Yang H-I, Chen WJ, Su M-W, Chu H-W, Shen C-Y. Taiwan Biobank: A rich biomedical research database of the Taiwanese population. Cell Genomics. 2022;2. doi: 10.1016/j.xgen.2022.100197

26. Dong Z, Guo S, Yang Y, Wu J, Guan M, Zou H, Jin L, Wang J. Association between ABCG 2 Q141K polymorphism and gout risk affected by ethnicity and gender: a systematic review and meta-analysis. International journal of rheumatic diseases. 2015;18:382–391. doi: 10.1111/1756-185X.12519

27. Zhang D, Ding Y, Wang X, Xin W, Du W, Chen W, Zhang X, Li P. Effects of ABCG2 and SLCO1B1 gene variants on inflammation markers in patients with hypercholesterolemia and diabetes mellitus treated with rosuvastatin. European journal of clinical pharmacology. 2020;76:939–946. doi: 10.1007/s00228-020-02882-4

28. Cleophas M, Joosten L, Stamp LK, Dalbeth N, Woodward OM, Merriman TR. ABCG2 polymorphisms in gout: insights into disease susceptibility and treatment approaches. Pharmacogenomics and personalized medicine. 2017:129–142. doi: 10.2147/PGPM.S105854

29. Kim H, Song T-J, Yee J, Kim D-H, Park J, Gwak HS. ABCG2 gene polymorphisms may affect the bleeding risk in patients on apixaban and rivaroxaban. *Drug Design*, Development and Therapy. 2023:2513–2522. doi: 10.2147/DDDT.S417096

30. Nakagawa J, Kinjo T, Aiuchi N, Ueno K, Tomita H, Niioka T. Impacts of pregnane X receptor and cytochrome P450 oxidoreductase gene polymorphisms on trough concentrations of apixaban in patients with non-valvular atrial fibrillation. European Journal of Clinical Pharmacology. 2023;79:127–135. doi: 10.1007/s00228-022-03424-w

31. Wang Z, Li Y, He X, Fu Y, Li Y, Zhou X, Dong Z. In vivo evaluation of the pharmacokinetic interactions between almonertinib and rivaroxaban, almonertinib and apixaban. Frontiers in Pharmacology. 2023;14:1263975. doi: 10.3389/fphar.2023.1263975

32. Gulilat M, Keller D, Linton B, Pananos AD, Lizotte D, Dresser GK, Alfonsi J, Tirona RG, Kim RB, Schwarz UI. Drug interactions and pharmacogenetic factors contribute to variation in apixaban concentration in atrial fibrillation patients in routine care. Journal of Thrombosis and Thrombolysis. 2020;49:294–303. doi: 10.1007/s11239-019-01962-2

33. Ueshima S, Hira D, Kimura Y, Fujii R, Tomitsuka C, Yamane T, Tabuchi Y, Ozawa T, Itoh H, Ohno S. Population pharmacokinetics and pharmacogenomics of apixaban in Japanese adult patients with atrial fibrillation. British journal of clinical pharmacology. 2018;84:1301–1312. doi: 10.1111/bcp.13561

34. Ueshima S, Hira D, Fujii R, Kimura Y, Tomitsuka C, Yamane T, Tabuchi Y, Ozawa T, Itoh H, Horie M. Impact of ABCB1, ABCG2, and CYP3A5 polymorphisms on plasma trough concentrations of apixaban in Japanese patients with atrial fibrillation. Pharmacogenetics and genomics. 2017;27:329-336. doi: 10.1097/FPC.0000000000000294

35. Nakagawa J, Kinjo T, Iizuka M, Ueno K, Tomita H, Niioka T. Impact of gene polymorphisms in drug-metabolizing enzymes and transporters on trough concentrations of rivaroxaban in patients with atrial fibrillation. Basic & Clinical Pharmacology & Toxicology. 2021;128:297–304. doi: 10.1111/bcpt.13488

36. Cheng S-T, Wu S, Su C-W, Teng M-S, Hsu L-A, Ko Y-L. Association of ABCG2 rs2231142-A allele and serum uric acid levels in male and obese individuals in a Han Taiwanese population. Journal of the Formosan Medical Association. 2017;116:18–23. doi: 10.1016/j.jfma.2015.12.002

37. Kanbay M, Segal M, Afsar B, Kang D-H, Rodriguez-Iturbe B, Johnson RJ. The role of uric acid in the pathogenesis of human cardiovascular disease. Heart. 2013;99:759–766. doi: 10.1136/heartjnl-2012-302535

38. Liu J, Yang W, Li Y, Wei Z, Dan X. ABCG2 rs2231142 variant in hyperuricemia is modified by SLC2A9 and SLC22A12 polymorphisms and cardiovascular risk factors in an elderly community-dwelling population. BMC Medical Genetics. 2020;21:1–9. doi: 10.1186/s12881-020-0987-4

39. Chen JH, Chuang SY, Chen HJ, Yeh WT, Pan WH. Serum uric acid level as an independent risk factor for all-cause, cardiovascular, and ischemic stroke mortality: a Chinese cohort study. Arthritis Care & Research. 2009;61:225–232. doi: 10.1002/art.24164

40. Kolz M, Johnson T, Sanna S, Teumer A, Vitart V, Perola M, Mangino M, Albrecht E, Wallace C, Farrall M. Meta-analysis of 28,141 individuals identifies common variants within five new loci that influence uric acid concentrations. PLoS genetics. 2009;5:e1000504. doi: 10.1371/journal.pgen.1000504

41. Fu S, Luo L, Ye P, Xiao W. Epidemiological associations between hyperuricemia and cardiometabolic risk factors: a comprehensive study from Chinese community. BMC cardiovascular disorders. 2015;15:1–5. doi: 10.1186/s12872-015-0116-z

42. Ee PLR, Kamalakaran S, Tonetti D, He X, Ross DD, Beck WT. Identification of a novel estrogen response element in the breast cancer resistance protein (ABCG2) gene. Cancer research. 2004;64:1247–1251. doi: 10.1158/0008-5472.can-03-3583

43. Chang F-W, Fan H-C, Liu J-M, Fan T-P, Jing J, Yang C-L, Hsu R-J. Estrogen enhances the expression of the multidrug transporter gene ABCG2—increasing drug resistance of breast cancer cells through estrogen receptors. International Journal of Molecular Sciences. 2017;18:163. doi: 10.3390/ijms18010163

44. Weng L-C, Choi SH, Klarin D, Smith JG, Loh P-R, Chaffin M, Roselli C, Hulme OL, Lunetta KL, Dupuis J. Heritability of atrial fibrillation. Circulation: Cardiovascular Genetics. 2017;10:e001838. doi: 10.1161/CIRCGENETICS.117.001838

45. Gudbjartsson DF, Arnar DO, Helgadottir A, Gretarsdottir S, Holm H, Sigurdsson A, Jonasdottir A, Baker A, Thorleifsson G, Kristjansson K. Variants conferring risk of atrial fibrillation on chromosome 4q25. Nature. 2007;448:353–357. doi: 10.1038/nature06007

46. Tucker NR, Clauss S, Ellinor PT. Common variation in atrial fibrillation: navigating the path from genetic association to mechanism. Cardiovascular research. 2016;109:493–501. doi: 10.1093/cvr/cvv283

47. Maharani N, Ting YK, Cheng J, Hasegawa A, Kurata Y, Li P, Nakayama Y, Ninomiya H, Ikeda N, Morikawa K. Molecular mechanisms underlying urate-induced enhancement of Kv1. 5 channel expression in HL-1 atrial myocytes. Circulation Journal. 2015;79:2659-2668. doi: 10.1253/circj.CJ-15-0416

48. Doshi M, Takiue Y, Saito H, Hosoyamada M. The increased protein level of URAT1 was observed in obesity/metabolic syndrome model mice. *Nucleosides*, Nucleotides and Nucleic Acids. 2011;30:1290–1294. doi: 10.1080/15257770.2011.603711

49. Alrajeh K, Roman YM. The frequency of rs2231142 in ABCG2 among Asian subgroups: implications for personalized rosuvastatin dosing. Pharmacogenomics. 2023;24:15–26. doi: 10.2217/pgs-2022-0155

50. Abdrakhmanov A, Akilzhanova A, Shaimerdinova A, Zhalbinova M, Tuyakova G, Abildinova S, Albayev R, Ainabekova B, Chinybayeva A, Suleimen Z. The distribution of the genotypes of ABCB1 and CES1 polymorphisms in Kazakhstani patients with atrial fibrillation treated with DOAC. Genes. 2023;14:1192.

